# Awareness and perceptions among members of a Japanese cancer patient advocacy group concerning the financial relationships between the pharmaceutical industry and physicians: a mixed-methods analysis of survey data

**DOI:** 10.1101/2021.06.26.21259442

**Authors:** Anju Murayama, Yuki Senoo, Kayo Harada, Yasuhiro Kotera, Hiroaki Saito, Toyoaki Sawano, Yosuke Suzuki, Tetsuya Tanimoto, Akihiko Ozaki

**Affiliations:** Medical Governance Research Institute, Minato-ku, Tokyo, Japan; Human Sciences Research Centre, University of Derby, Derby, Derbyshire, United Kingdom; Department of Gastroenterology, Sendai Kousei Hospital, Sendai, Miyagi, Japan; Department of Surgery, Jyoban Hospital of Tokiwa Foundation, Iwaki, Fukushima, Japan; Department of Internal Medicine, Navitas Clinic Kawasaki, Tokyo, Japan; Department of Breast Surgery, Jyoban Hospital of Tokiwa Foundation, Iwaki, Fukushima, Japan

**Keywords:** conflict of interest, ethics, Japan, financial relationship, patient-centered care, pharmaceutical industry

## Abstract

**Objectives:** Financial conflicts of interest (FCOI) between pharmaceutical companies (Pharma) and healthcare domains may unduly influence physician-led clinical practice and patient-centered care. However, the extent of awareness and perceptions of FCOI among Japanese cancer patients remains unclear. This study aimed to assess these factors and their impacts on physician trustworthiness among Japanese cancer patients.

**Methods:** A cross-sectional study using self-administered surveys was conducted on a Japanese cancer patient advocacy group with 800 registered members from January to February 2019. Main outcome measures included awareness and perceptions of physician-Pharma interactions, their impact on physician trustworthiness, and attitudes towards FCOI among professions. We also performed thematic analyses on additional comments responders provided in the surveys.

**Results:** Among the 524 invited members, 96 (18.3%) completed the questionnaire. Of these, 69 (77.5%) were cancer patients. The proportion of participants aware of such interactions ranged from 2.1% to 65.3%, depending on the interaction type. Participants were generally neutral on how the interactions would affect physician trustworthiness. A large proportion of participants agreed that these interactions were unethical, could influence physicians’ prescribing behavior leading to unnecessary prescriptions, and negatively affect physician trustworthiness. Qualitative responses (n=56) indicated that patients expected physicians to use sound ethical judgment and avoid accepting incentives. Participants were also concerned about their treatment and the undue influence of FCOI on physicians.

**Conclusion:** Most participants were aware of at least one FCOI between Pharma and physicians and perceived them negatively. Further efforts to regulate FCOI appear necessary to protect patient-centered care.

## Main text

### Introduction

In recent years, patient-centered care has widely been considered a key component of healthcare delivery. Defined as one of the Six Domains of Healthcare Quality introduced by the National Academy of Medicine,[1] patient-centered care focuses on delivering healthcare respecting patient preferences, needs, and values. Therefore, financial conflicts of interest (FCOI) between pharmaceutical companies (Pharma) and healthcare domains are of significant concern, mainly due to potential bias in patient care and medical institutions’ operation.[2–6] Key criteria of patient-centered care include full transparency and fast delivery of information.[7–9] In the world today, there are transparency initiatives such as the Sunshine Act and Open Payments Database in the United States (US),[10–12] Transparency in Healthcare in France,[5] Euro for Doctors and LeitlinienWatch in Germany,[13, 14] Disclosure UK in the United Kingdom,[15–17] Disclosure Australia in Australia,[18, 19] and Money Database in Japan,[2, 20, 21], all of which emphasize improving transparency and raising awareness of FCOI among patients and the general public to achieve excellent patient-centered care.

However, there remains an ongoing controversy about whether these transparency initiatives improve the general awareness of FCOI.[10, 12, 22] A previous qualitative study conducted before these initiatives reported that most cancer clinical trial participants had a positive or neutral view of physicians receiving research funding from Pharma.[23] Moreover, there has been little improvement in awareness of FCOI even after the launch of transparency initiatives.[10, 22, 24] Rather, it has been reported that public trust in healthcare professionals dropped following the launch of these initiatives.[25] These findings demonstrate the limited effect on the awareness of FCOI by these transparency initiatives.

Nonetheless, recent discussions on this issue have not considered specific populations with a higher interest in this topic, including patients with critical illnesses or their caregivers. Among patient populations, cancer patients are particularly important, given the critical nature of the disease and its burden,[26] as well as the development of numerous novel and expensive therapeutics.[27, 28] Since novel anticancer drugs are potentially highly profitable, these agents’ development remains among Pharma’s highest priorities.[29] Therefore, these findings suggest the importance for cancer patients to understand FCOI among Pharma and healthcare domains.

The issues surrounding FCOI are of particular relevance in Japan due to its universal health coverage and the fact that its pharmaceutical market is the third-largest globally, after the US and China.[30] In 2018, anticancer drugs’ annual pharmaceutical sales exceeded 1.24 trillion Japanese yen (JPY; 11 billion US dollars [USD]), accounting for 12% of Japan’s total annual pharmaceutical sales. Despite an overall decline in Japan’s pharmaceutical sales, the oncology drug market continues to expand and is projected to exceed 13 billion USD by 2025.[31] There also has been progress in terms of Japanese transparency initiatives. Along with several non-profit organizations, the Japan Pharmaceutical Manufacturer Association has, since it first developed transparency guidelines in 2011,[32] led efforts to improve transparency in Pharma and healthcare domains’ financial relationships. For instance, since 2013, all Japan Pharmaceutical Manufacturer Association members have voluntarily disclosed payments to physicians on their respective websites following established transparency guidelines.[32] Also, Japanese non-profit organizations, including the Medical Governance Research Institute and Waseda Chronicle, have developed the Money Database. This database enables the general public to learn about financial relationships among individual pharmaceutical companies and healthcare sectors.[2, 33–36] While these organizations disclose information regarding industry payments made to physicians, how Japanese cancer patients perceive the information remains unclear.

In sum, the present study’s purpose was to examine awareness of physician-Pharma interactions and their impact on physicians’ trust while also appraising perceptions of these payments among Japanese cancer patients.

## Methods

### Study setting, design, and participants

The study population consisted of 524 individuals of the Cancer Treatment Society for Citizens, a support group for cancer patients established in 2004 with 800 registered members. In response to increasing trends towards second opinions in oncology,[37] this support group aids patients in determining the most appropriate treatment by offering referrals with radiation oncologists certified by both the Japan Radiology Society and the Japanese Society for Radiation Oncology. Along with the referral service for second opinions, the Cancer Treatment Society for Citizens also offers educational programs and publishes articles by cancer specialists on its website to promote general awareness of contemporary cancer research and treatment options.

### Data collection

Data collection occurred from January 9 to February 10, 2019, using the structured questionnaire. The printed survey questionnaire was distributed to all 524 Cancer Treatment Society members for Citizens registered on the society’s mailing list with the society’s quarterly newsletter on January 9, 2019. Members that agreed to participate in the study completed the questionnaire and returned it, as requested, via the enclosed self-addressed, stamped envelope by February 10, 2019. To mitigate the response biases, the authors were not directly involved in the survey distribution or collection.

### Survey sheet

We compiled our cross-sectional questionnaire survey, taking into account previous studies and the local context of Japanese cancer care.[38, 39] The survey included 51 questions covering the following items: 1) the status of disease progression and demographic characteristics such as gender, education level, and medical history; 2) awareness of the physician-Pharma interactions, including physician receipt of gifts, payments and research rewards from Pharma (e.g., *“Do you know that some physicians would receive pamphlets or leaflets concerning products manufactured by pharmaceutical companies from their sales representatives?”*) on a three-point Likert scale (*Yes, No, or Not sure*); 3) influence of the interactions’ on trust (e.g., *“How would your trust in your physician be influenced if your physician received pamphlet or leaflet with information about the products from a pharmaceutical sales representative?”*) on a five-point Likert scale; 4) perception of trust and regulation on the interactions (e.g., *“Stricter regulations about gifts, meals, and honoraria from pharmaceutical companies to physicians are needed.”*) on a five-point Likert scale; 5) attitude towards the industrial payments across professions (e.g., *“It is problematic for physicians to receive gifts, meals, and other entertainment from pharmaceutical sales representatives.”*) on a five-point Likert scale; and 6) an open-ended question about participants’ perception on the FCOI between Pharma and physicians (“*Please freely describe your thoughts about non-research-related offerings from Pharma to a physician (e.g., gifts, free meals, monetary incentives for a lecture)*”). The survey questions are described in greater detail in the Supplementary Methods (Supplementary Material 1).

### Data analysis

The initial descriptive analysis included the patients’ sociodemographic and clinical variables and all questions concerning participant awareness, influence on trust in physicians, perceptions, and attitudes towards FCOI.

Next, to assess the association between awareness (outcome variable 1) and the impact of physician-Pharma interaction on physicians’ trustworthiness (outcome variable 2) and each participant’s sociodemographic and clinical factors, we constructed logistic regression models for trust in physicians. We re-categorized the following variables as appropriate and set them as predictor variables: annual income, highest educational qualification, medical history, type of business, hospital types, cancer stage, year of diagnosis, experience with cancer recurrence, experience with pharmacotherapy, experience with radiotherapy, and experience with surgical treatments. Further in the logistic analysis, to assess the factors related to participant awareness and to ensure statistical stability, the awareness status was re-categorized into two groups with the outcome variable set as those who responded “*Yes*” to one or more of the 11 questions about awareness, and those who responded *“No”* or *“Not sure”* to all questions. Similarly, the relationship between influence on physicians’ trust and the predictor variables was also assessed using logistic regression analysis. In the analysis of the influence on trust in physicians, the variables measuring physicians’ trust were aggregated into two types, with the outcome variable set as those who reported “*decreased trust in physicians*” or “*slightly decreased trust in physicians*” on at least one question, and the other respondents. In both analyses, multicollinearity was of no concern (VIFs < 10).

These data analyses were informed by similar previous studies reported by Ammous et al. and Green et al.[38, 40] All analyses, including descriptive statistics, were performed using Stata version 15 (STATA Corp., College Station, TX, United States). Conversion of JPY to USD used the 2019 average monthly exchange rate of 109.0 JPY per USD.

Lastly, participants’ responses to the open-ended questions regarding perception on FCOI were analysed thematically, following Braun and Clarke.[41]

### Ethics approval

The Institutional Review Board of Medical Governance Research Institute granted ethics approval of this study (MG2018-07-0928), adhering to the Ministry of Health Labour and Welfare and The Ministry of Education guidelines, Culture, Sports, Science, and Technology in Japan.

## Results

### Participants

All surveys returned by February 10, 2019, were considered. Of the 524 eligible survey participants, 96 completed the questionnaire (completion rate=18.3%). Table 1 presents participant sociodemographic and clinical characteristics. Respondents were predominantly male (67.7%, 63/93), older than 70 years (52.3%, 49/94), and 53.8% (50/93) had educational attainment of a bachelor’s degree or higher. Further, 55.9% (52/93) were unemployed, and about half (46.5%, 40) had an annual household income of over $36,697, roughly the average of Japanese households in 2018. Of the 89 respondents providing primary disease status, 69 (77.5%) were cancer patients, and 20 (22.5%) were non-cancer patients (e.g., family members of cancer patients).

**Table 1.** Demographic features of the study population.

| Variables |  |
| --- | --- |
| **Gender, N (%)** |  |
| Male | 63 (67.7) |
| Female | 30 (32.3) |
| Missing | 3 |
| **Age category, N (%)** |  |
| ≤60 | 15 (16.0) |
| 61-70 | 30 (31.9) |
| 71-80 | 38 (40.4) |
| 81-90 | 11 (11.7) |
| Missing | 2 |
| **Annual family income, N (%)** |  |
| < 2 million JPY (< 18,349 USD) | 6 (7.0) |
| 2-4 million JPY (18,349-36,697 USD) | 40 (46.5) |
| 4-6 million JPY (36,697-55,046 USD) | 14 (16.3) |
| 6-8 million JPY (55,046-73,394 USD) | 12 (14.0) |
| 8-10 million JPY (73,394-91,743 USD) | 5 (5.8) |
| >10 million JPY (>91,743 USD) | 9 (10.5) |
| Missing | 10 |
| **Type of business, N (%)** |  |
| Unemployed | 52 (55.9) |
| Self-employed | 15 (16.1) |
| Full-time job | 11 (11.8) |
| Part-time job | 7 (7.5) |
| Other | 8 (8.6) |
| Missing | 3 |
| **Educational background, N (%)** |  |
| Less than high school graduate | 1 (1.1) |
| High school graduate | 27 (29.0) |
| Associate degree or diploma | 15 (16.1) |
| Bachelor's degree or more<br>(bachelor, master and doctoral degree) | 50 (53.8) |
| Missing | 3 |
| **Type of cancer, N (%)** |  |
| Not cancer patient | 20 (22.5) |
| Prostate cancer | 24 (27.0) |
| Lung cancer | 5 (5.6) |
| Breast cancer | 5 (5.6) |
| Colorectal cancer | 4 (4.5) |
| Gastric cancer | 4 (4.5) |
| Other type of cancer | 27 (30.3) |
| Missing | 7 |
| **Cancer's stage, N (%)\*** |  |
| Stage 1 | 23 (35.9) |
| Stage 2 | 16 (25.0) |
| Stage 3 | 10 (15.6) |
| Stage 4 | 5 (7.7) |
| Unclear | 10 (15.6) |
| Missing | 5 |
| **Year of diagnosis, N (%)\*** |  |
| 2018 | 4 (5.8) |
| 2017 | 6 (8.7) |
| 2016 | 3 (4.4) |
| 2015 | 4 (5.8) |
| Before 2015 | 52 (75.4) |
| **Hospital, N (%)\*** |  |
| Cancer hospital | 20 (29.4) |
| National university | 9 (13.2) |
| Private university | 9 (13.2) |
| National municipal hospital | 10 (14.7) |
| Private municipal hospital | 11 (16.2) |
| Other hospital | 9 (13.2) |
| Missing | 1 |
| **Previous cancer recurrence\*** |  |
| Yes | 13 (20.0) |
| No | 47 (72.3) |
| Not clear | 5 (7.7) |
| Missing | 4 |
| **Count of previous cancer reoccurrence, N (%)** |  |
| 1 | 6 (42.9) |
| 2 | 2 (14.3) |
| 3 | 1 (7.1) |
| 6 | 1 (7.1) |
| 7 | 1 (7.1) |
| 10 | 1 (7.1) |
| Unknown | 2 (14.3) |
| **Treatment which you have ever had, N (%)\*, \*\*** |  |
| Anticancer drug | 22 (16.5) |
| Molecularly targeted drug | 5 (3.8) |
| Hormone therapy | 23 (17.3) |
| Radiation therapy | 37 (27.8) |
| Surgery | 39 (29.3) |
| Other | 4 (3.0) |
| Never | 3 (2.3) |
| **Treatment which you have now, N (%)\*\* \*\*** |  |
| Anticancer drug | 2 (2.9) |
| Molecularly targeted drug | 2 (2.9) |
| Hormone therapy | 10 (14.5) |
| Radiation therapy | 2 (2.9) |
| Other | 11 (15.9) |
| Not having | 42 (60.9) |

\* The denominator was the number of people with any type of cancer. (n=69)
\*\* Multiple answers were possible.
Abbreviations: JPY; Japanese yen, USD; US dollar

Of the 69 participants self-identifying as cancer patients, 75.4% (52/64) received their diagnosis before 2015. Further, 30.9% (21/68) of these participants primarily received treatment at a municipal hospital, while 60.9% (42/69) did not actively receive any treatment during the survey period. Lastly, 39 (56.5%) reported previous pharmaceutical treatments for cancer, including anticancer drugs, molecularly targeted drugs, or hormone therapy.

## Quantitative findings

### Awareness of physician-Pharma interactions

Figure 1 presents the participants’ awareness of physician-Pharma interactions. The proportion of the participants aware of these interactions ranged from 2.1% to 65.3%, depending on the type of interaction. The interaction with the largest proportion of participant awareness was pamphlets and leaflets (65.3%), followed by stationery (64.2%) and free drug samples (53.1%). In contrast, the participants were the least aware of possession of stocks (2.1%). Although Japanese companies belonging to the Japan Pharmaceutical Manufacturer Association have disclosed payments to healthcare sectors since 2013, only 10.5% (10/95) of the survey responders were aware of such disclosure. Overall, 80.2% (77/96) of participants were aware of at least one physician-Pharma interaction. Detailed results of awareness by participant demographic characteristics are shown in Supplementary Material 5. None of the sociodemographic and clinical variables in the logistic regression analysis significantly predicted awareness of physician-Pharma interactions (*p* ≧ 0.05) (Supplementary Material 6).

**Figure 1.**
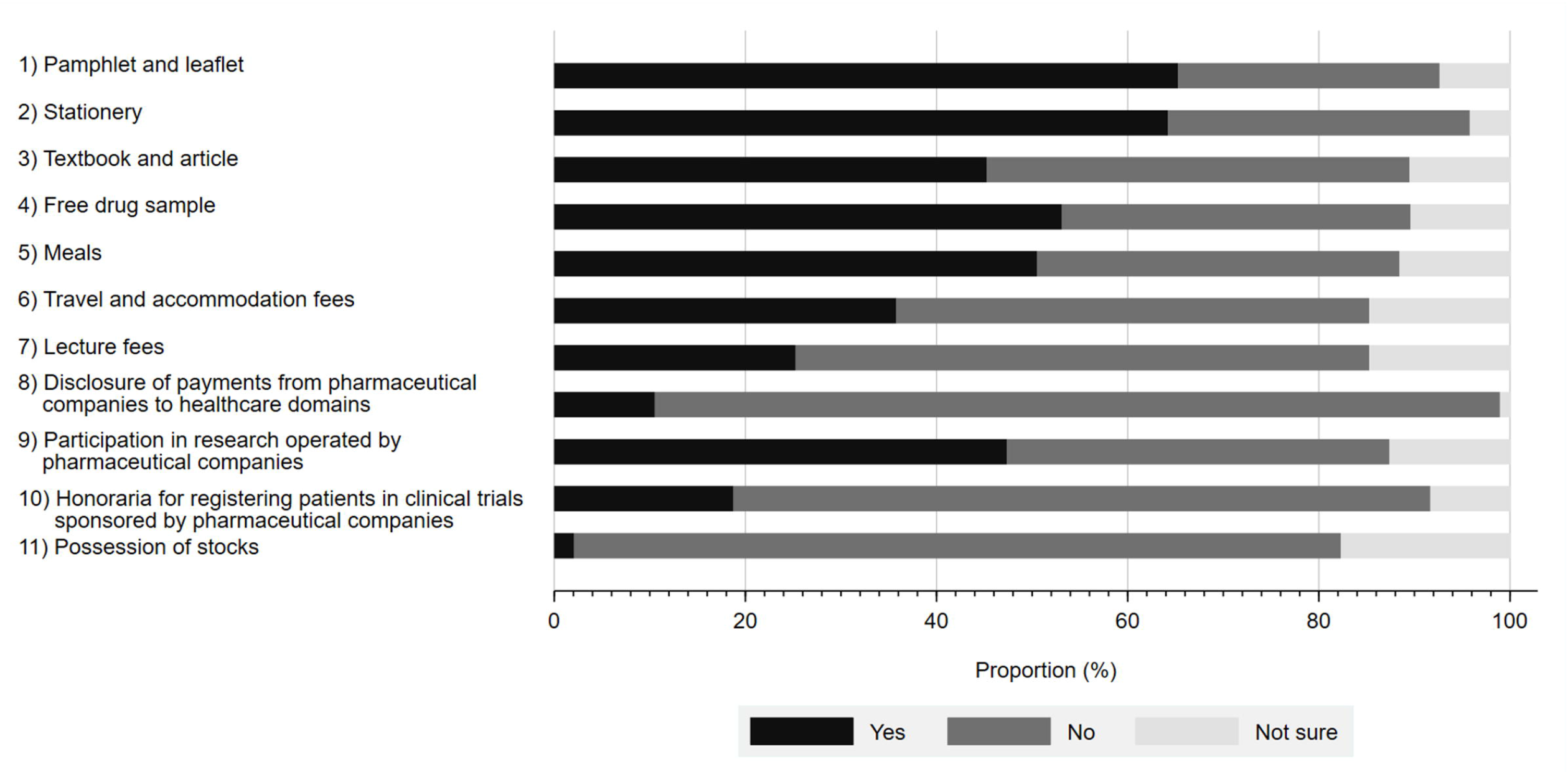
Participants’ awareness of interactions between physicians and pharmaceutical companies.

### Influence of physician-Pharma interactions on trust in physicians

Figure 2 shows the impact of physician-Pharma interactions on patients’ trust in physicians. Although participants overwhelmingly responded that most physician-Pharma interactions neither increased nor decreased their trust in physicians, 81.2% (58.3% decrease in trust, 22.9% slightly decreased trust) agreed that stock ownership would negatively impact their trust in physicians. Additionally, accepting either honoraria for registering patients in industry-sponsored research (52.1% decrease in trust, 26.0% slightly decreased trust) or lecture fees (31.6% decrease in trust, 30.5% slightly decreased trust) also substantially impacted participant perceptions. Interestingly, few participants reported that indirect gifts, such as accepting free samples of prescription drugs (12.8% decrease in trust, 25.5% slightly decreased trust), would reduce trust in physicians. On the other hand, participation in industry-sponsored research generally appeared to increase physician trustfulness by 13.6% (increased trust 2.1%, slightly increased trust 11.5%) among participants. Overall, 90.6% of participants marked decreased trust or slightly decreased trust on at least one question. Associations between participant demographic characteristics and trust in physicians are shown in Supplementary Material 7. Our regression analysis revealed that none of the sociodemographic or clinical variables significantly predicted whether patients’ trust in physicians would decrease or not (*p* ≧ 0.05) (Supplementary Material 8).

**Figure 2.**
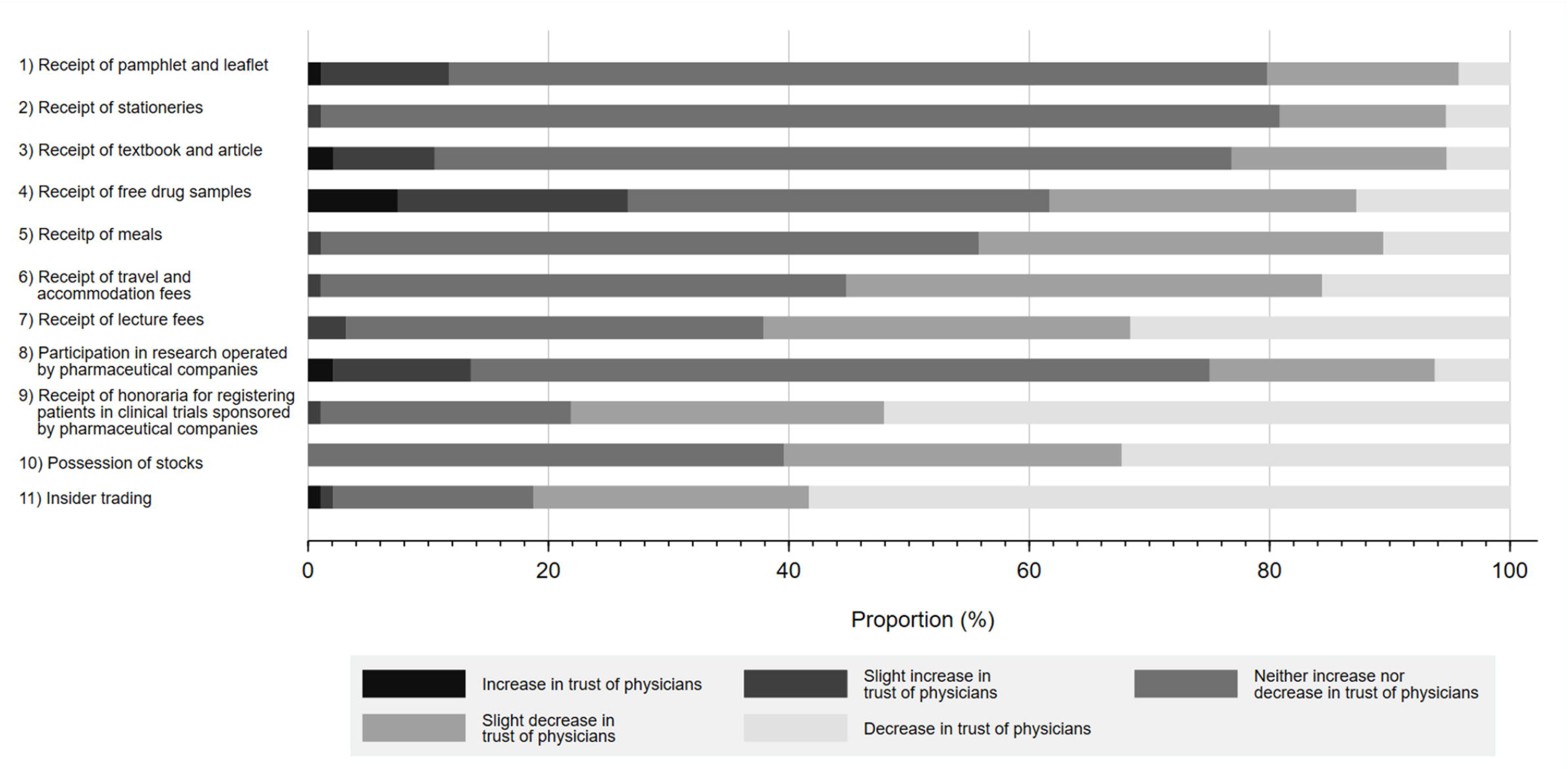
Influence of physician-Pharma interactions on trust in physicians.

### Perception on physician-Pharma interactions

Figure 3 details the reported perceptions on a series of statements regarding physician-Pharma interactions and regulations associated with each. Overall, a substantial proportion of participants either agreed or slightly agreed that gifts from pharmaceutical companies have a significant influence on physicians’ prescription behavior (35.8% agree, 38.9% slightly agree). A similar proportion viewed such gifts as unethical (31.6% agree, 30.5% slightly agree), contributing to unnecessary prescriptions (34.7% agree, 37.9% slightly agree), and negatively influencing patients’ trust in physicians (38.9% agree, 31.6% slightly agree). And while many participants acknowledged the need to regulate physician-Pharma interactions, more concluded that there should be greater self-regulation by industry (60.0% agree, 26.3% slightly agree) or physicians (60.2% agree 24.7% slightly agree) as opposed to legal regulation (45.3% agree, 29.5% slightly agree).

**Figure 3.**
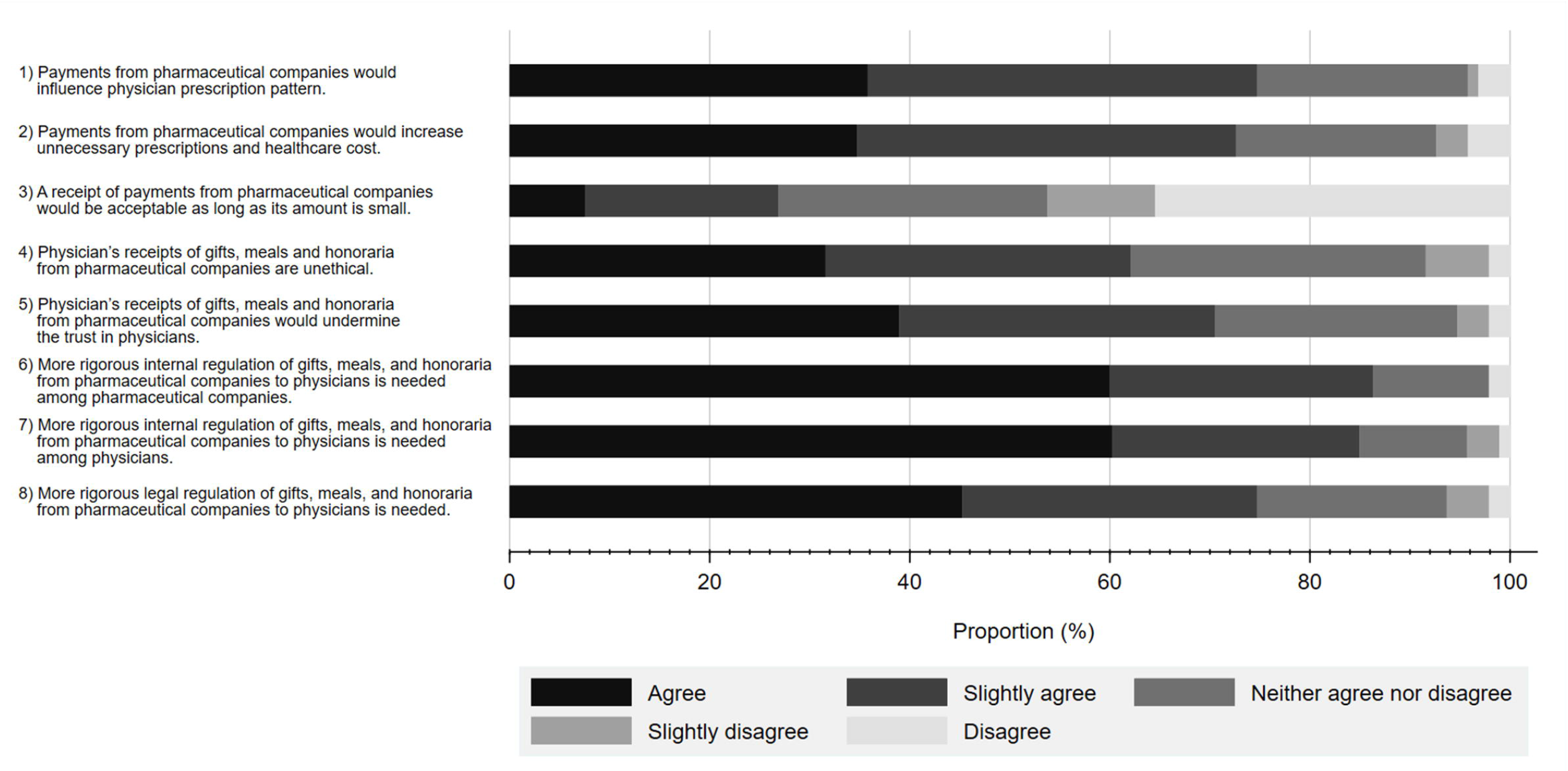
Participants’ perceptions on statements regarding interactions between physicians and pharmaceutical companies and regulations associated with them.

Figure 4 shows participants’ perceptions on acceptable amounts and frequency of non-research payments from Pharma to physicians. In total, 75.9% of the participants considered an interaction worth 10,000 JPY (91.7 USD) or below as acceptable, and 92.8% believed that the annual amount should be less than 1 million JPY (9,174 USD). Further, 77.8% of the participants considered one interaction every few months as acceptable.

**Figure 4.**
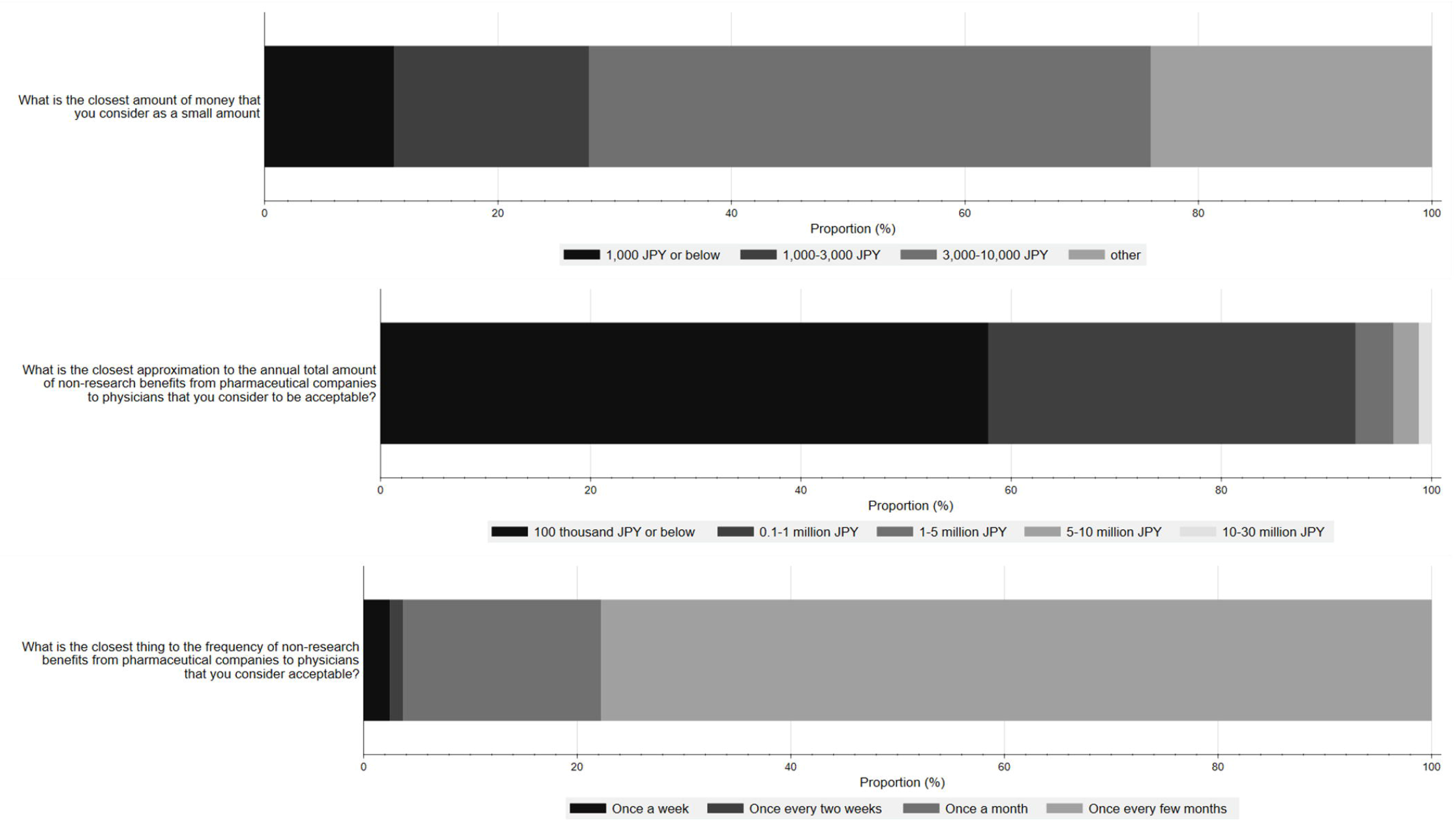
Participants’ perception of acceptable amounts and frequency of non-research payments from pharmaceutical companies to physicians.

### Attitude towards various professional FCOI

Figure 5 shows the participants’ perceptions towards potential FCOI in various professions, including court judges, referees, politicians, physicians, and business professionals. Overall, a larger percentage of participants considered it problematic for court judges, politicians, or referees to receive gifts and meals from lawyers (agree 92.6%, slightly agree 6.3%), lobbyists (agree 75.5%, slightly agree 20.2%), or players (agree 89.4%, slightly agree 4.3%), respectively, than for physicians to accept gifts from Pharma (agree 58.5%, if slightly agree 23.3%).

**Figure 5.**
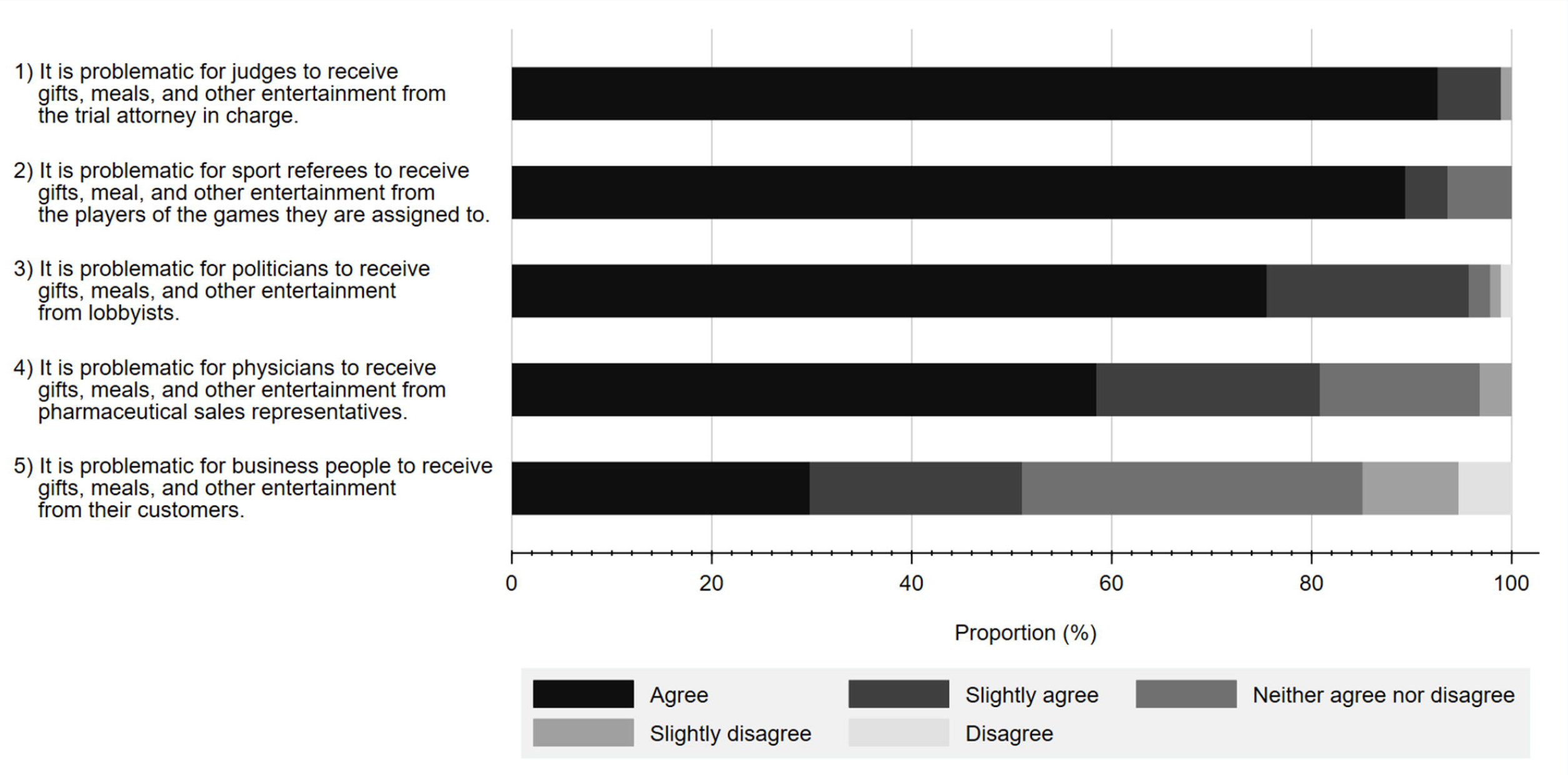
Participant attitude about conflicts of interest among various professionals.

## Qualitative findings

### Open-ended responses

A thematic analysis of the survey’s open-ended question, responded to by 56 (58.3%) of the participants, identified four themes: 1) Perception towards the FCOI, 2) Concerns about the participant’s treatment, 3) Reasons for physician-Pharma interactions and 4) Possible solutions from the patient’s perspective.

#### Theme 1: Perception towards the FCOI

The majority of respondents agreed that physicians should not receive honoraria except for research purposes. However, even the receipt of honoraria for research purposes was conditioned on the premise that such awards would not interfere with physicians’ ability to deliver high-quality patient-centered care.

> “Never permit benefits except for research purposes. Pharmaceutical companies, including those who are involved in research, should realize that the pharmaceutical companies are responsible for the lives of people. We don’t want doctors to accept one penny of honorarium.”

Further, concerning lecture fees, a few participants appeared to consider that physicians deserve to receive payment to some extent due to the physicians’ efforts in preparing for the lecture.

> “Physicians are busy and time-restricted, so I think it’s reasonable for them to receive it (a lecture fee). But I disagree with any other kind of benefit-sharing between them, because that may hinder optimal selection of the prescription.”

The majority of respondents wanted the financial transactions to be minimized, except for reasonable research payments. This is in line with the results from our quantitative analysis. For example, more than half of the participants considered non-research benefits of less than 100,000 JPY (917 USD) per year to be appropriate.

#### Theme 2: Concerns about the participant’s treatment

Many participants expressed concerns about the influence of physician-Pharma interactions on treatment decisions and the treatment they receive. Participants mostly wanted doctors to treat patients based on their authentic judgment and not be influenced by these interactions.

> “Doctors are also human, and if they are incentivized, then they may have to make a judgment that is favored by a pharmaceutical company. I’ve heard of such things, and I think doctors should put their patients first, as cliché as it may sound. Patients trust their doctors and put their lives in doctor’s hands. I hope that not all doctors are in favor of Pharma.”

> “My former doctor used to go to ‘study meetings’ a lot. To treat my advanced cancer, this doctor strongly recommended a medicine that he had just learned about in these study meetings. Although I trusted him to treat me, having learned about the financial relationships between physicians and pharmaceutical industry in this research, now I would have made different decisions about his recommendations.”

In alignment with our quantitative analysis results, many participants expressed concern about the influence of physician-Pharma interactions on physician’s prescription decisions in general. Additionally, several participants voiced unease about their physicians and the treatments they received.

#### Theme 3: Reasons of physician-Pharma interactions

Many participants concluded that a lack of physician ethical norms could cause FCOI between Pharma and healthcare sectors.

> “I think it comes down to the ethics and morality of each physician.”

Besides physicians’ morality, two participants raised the current government policy for science as an ongoing issue of physician-Pharma interactions. They indicated that the decline in the national budget for scientific research led to the financial dependence on Pharma.

> “I think there is also a problem with the way the Japanese government funds basic research, alongside physician ethics and morality, if we want to ensure that (healthcare) is not controlled by pharmaceutical companies.”

In contrast to Themes 1 and 2, neither of these two potential reasons were clarified in the quantitative results.

#### Theme 4: Possible solutions from the patient perspective

The open-ended responses about possible solutions for FCOI comprised three main positions: improving public awareness of physician-Pharma interactions, strengthening legal or public FCOI regulation, and educating healthcare professionals on ethical norms.

Several participants suggested possible solutions for the FCOI between Pharma and healthcare sectors. Improving public awareness of physician-Pharma interactions was the idea suggested most often.

Also, regulating the FCOI by professional associations or law was suggested by several participants while also acknowledging the need for receipts for transport, meals, and honoraria relating to lectures and research. These comments also supported the quantitative results that more than three-fourths of the participants agreed with some form of regulation.

> “If [doctors] give a lecture, [doctors] may get transportation, food, and honorarium. It is generally an acceptable amount of money. If the basis on which pharmaceutical companies decide the concrete amount of the payments to physicians is unclear, the criteria should be decided by the professional association or academic society, taking into account the age and experience of the speaker.”

On the other hand, a few participants insisted a regulatory regime for physician-Pharma interactions based on each physician’s morality rather than on legal or public regulation. This would further indicate the need for the education of physicians on the perspective of ethical norms.

## Discussion

In this study, we investigated the awareness and perceptions of FCOI between Pharma and physicians among members of a Japanese cancer patient advocacy group demographically representative of the general Japanese cancer patient population.[42] We primarily found that most of the participants were aware of physician-Pharma interactions, although the extent of the awareness differed based on the nature of the interaction. We also found that participants mainly considered these interactions influential on clinical practice and agreed to the need for further regulation of physician-pharma interactions.

Although the awareness of the interactions in this study, such as receipt of stationeries (64.2%), travel fees (35.8%), lecture fees (25.3%), and disclosure of payments from Pharma to the healthcare sector (10.5%), were within the scope of previous studies (55–76%, 17–33.7%, 20–46%, 7.3–18.8%, respectively),[12, 22, 24, 38, 43, 44] the awareness of receiving drug samples (53.1%) was significantly lower than studies in the US (87–93.9%).[43, 45] In contrast, the awareness of physicians conducting research for Pharma (47.4%), receipt of meals (50.5%), and receipt of textbooks (45.3%) were higher than US and Lebanese studies (23–32%, 22–37%, 35–37%), respectively.[38, 40, 43] The underlying reasons for such differences among countries are not obvious. Still, stationaries and educational gifts, which could be present in clinicians’ offices, were more likely to be recognized than personal payments for lectures, travel, or accommodations, a trend which we observed previously.[43, 45–47]

One of the most interesting findings from this study was that despite ongoing disclosure efforts since 2013, only about 10% of participants knew major Japanese Pharma disclosed payments to physicians. Even so, the proportion of participants who were aware of such disclosures was within the range reported for previous studies conducted in the US (7.3%-18.8%).[12, 22, 24, 44] Nevertheless, this study’s participants may have had a more robust general interest in awareness of FCOI between Pharma and healthcare sectors. The qualitative study’s findings also revealed a lack of attentiveness on this issue, suggesting a need for proper countermeasure. Notably, during the study period on January 15, 2019, the Money Database was made publicly available in Japan. Since then, several academic papers and media reports have documented the financial relationships among Pharma and various healthcare sectors. [20, 34–36, 48–50] This potentially contributed to a recent increase in public attention to the issue of FCOI in Japan and warrants a follow-up study to further evaluate this issue.

This survey also demonstrated that most of the participants considered the presence of FCOI with Pharma as harming participants’ trust in physicians. Interestingly, although the results of the qualitative analyses indicated that a majority of respondents believed FCOI relating to research were acceptable when framed in the context of physicians participating in Pharma sponsored research, our quantitative findings noted a larger proportion of the participants (25%) reporting a decrease of trust in physicians than those who reported an increase of trust (13.6%). Further, 78.1% of the participants indicated a loss of trust when physicians received Pharma payments for participating in a clinical trial. This finding starkly contrasts with previous studies in other countries where the receipt of payments for lectures or conducting Pharma sponsored research were viewed as a positive symbol of good physicians by the public.[23, 51, 52]. Given that clinical trials can have positive consequences, including improving management strategies of specific conditions or expanding the treatment options of patients participating in clinical trials, this apparent skepticism of clinical trials among the Japanese public warrants attention. Nonetheless, FCOI between Pharma and physicians may cause bias in reporting research findings [53, 54] or medical scandals.[55] Thus, proper control of FCOI must accompany research involving Pharma.

In this study, more than three-quarters of participants considered general FCOI with Pharma to be influential in physician prescription in a certain way. The observed value (75.9%) was the highest in several previous studies reported in other countries, ranging from 29% in Turkey to 75.6% in Canada.[38, 43–45, 56, 57] Also, more participants agreed that physician-Pharma interactions would increase healthcare costs (72.6%) more so than other studies (33%–67.3%).[43] A possible reason for this may relate to the study population comprising cancer patients or their advocates. Essentially, this population is characterized by more significant health risks with few treatment alternatives outside of seeing a physician, [58–60] and associated regular and long-term care[61, 62], leading to greater familiarity and trust in their physicians.[58]

Aspects of Japanese culture likely also contribute to our findings. In particular, the Japanese medical profession has traditionally been held sacred, with physicians considered above reproach due to their ethos. Often more so than in other countries. Moreover, there exists a deep sense of shame and extreme disdain associated with improper behavior.[63, 64] Consequently, physicians with inappropriate financial entanglements may be seen as a kind of “defilement,” leading to distrust or disgust of such physicians by Japanese patients more strongly than among those in other countries.

More than 80% of participants answered that gifts accepted by physicians from Pharma were problematic. However, compared to other fields, fewer people considered accepting gifts in the medical field as more problematic than the legal, athletic, and political fields and less problematic compared to the general business world. This trend was similar to that observed in the US.[40] However, our study showed that more participants considered the physician-Pharma interactions to be unethical than their counterparts in the US (58.5% unethical and 22.3% somewhat unethical in our study versus 44% unethical and 15% somewhat unethical in the US).[40] Although we were unable to find an international comparison of physicians’ public perception related to ethical norms, our findings indicated that a greater percentage of the Japanese public might expect physicians to be ethical than the general public in other countries.

## Clinical implementation

In completing the questionnaires, participants provided several key perspectives in their quantitative and open-ended qualitative responses to properly improve patient-centered care and manage FCOI between Pharma and healthcare sectors.

As our previous studies revealed, the current framework for regulating FCOI between Pharma and healthcare sectors in Japan still has room for improvement, particularly regarding transparency.[20, 21, 35, 36, 65] Regarding FCOI, we observed among cancer patients a consensus that physicians, professional associations, and Pharma should exhibit higher ethical standards and self-regulate their FCOI rather than rely on government regulation. This perspective has important repercussions for future discussions on improving the transparency of FCOI in the Japanese healthcare system. One likely countermeasure would be developing voluntary disclosure databases initiated by professional organizations akin to Disclosure UK or Disclosure Australia.[9, 66] Another option would include the rigorous and legally binding FCOI disclosure standards established by the Sunshine Act and the Open Payment database in the US or the Transparency in Healthcare database in France.[5, 10–12]

Second, our survey elucidated that nearly 20% of participants were unaware of many kinds of physician-Pharma interactions despite representing a population with a high level of interest in medical issues. Given that numerous studies showed that FCOI between Pharma and healthcare sectors influence patient treatment [6, 67, 68] and that many patients wanted to know about these conflicts [12, 52, 69–71], it is noteworthy that patients could benefit significantly from knowledge about FCOI before choosing doctors and treatments.[44, 52, 72] Therefore, we suggest the possibility of exploring seminars focusing on direct communications with the general public to build stronger relationships between Pharma, healthcare sectors, and patients and to gain patient trust.[73] One study from Australia showed that small-sized workshops help improve patient knowledge about FCOI.[74]

In the meantime, the potential for increasing patient awareness of FCOI between Pharma and healthcare sectors may disrupt the long-standing, trusting relationship between patients and their physicians. As Kanter previously hypothesized,[72] our results suggest that patients with long-term and severe diseases like cancer would lose faith in their physicians over FCOI. Moreover, these concerns extended to a physician’s judgment of treatment options wherein some participants expressed similar concerns about the treatment they received. Specifically, this raises concerns about the general awareness of FCOI between Pharma and healthcare sectors, which could lead to some patients declining effective treatments suggested by their physicians. Therefore, careful consideration should be paid in communicating with patients about FCOI between Pharma and healthcare sectors.

Third, some participants suggested that there was a need to educate physicians about FCOI. According to Medical Professionalism in the New Millennium: A Physician Charter, maintaining trust by managing FCOI is one of the fundamental responsibilities for all physicians.[75] However, only one-third of Japanese medical schools undergo formal training on FCOI between Pharma and healthcare sectors.[76] Most physician-Pharma interactions are rooted in the first year of a physician’s training, after which physicians are continuously exposed to such interactions.[77, 78] For this reason, few physicians question these interactions or their influence on their clinical practice, and some physicians are not honest with their patients about FCOI.[76, 79–82] For example, more than 98% of Japanese medical students have financial interactions with Pharma.[83] Also, several studies revealed that junior physicians, more so than senior physicians, were more likely to accept interactions with Pharma[84, 85] and consider them appropriate and valuable.[85] Therefore, we would suggest that all medical schools establish a curriculum on FCOI addressing these relationships’ undue influence on clinical practice[6, 68, 86] and their impact on patient trust and care.[72] Given that even preclinical medical students can interact with Pharma, it might be advisable for this curriculum to be implemented early, by the second or third year in Japan.[76, 87, 88]

## Limitations

Our study has several limitations. First, it is worth noting that the response rate was relatively low at 18.3%. Many questions and the sensitive nature of the topic may have caused candidates to evade answering questions. The base response rate suggests that only candidates with a relatively strong interest in or problematic awareness of FCOI between Pharma and healthcare professionals may have participated in this study. Second, our study was conducted among the members of a cancer patient advocacy organization, many of whom have much more contact with Pharma and healthcare professionals than the general public and presumably greater interest in medical issues. Further, compared to Japan’s general cancer population in 2015, the current study population tended to be more male but with a similar age distribution.[42] Therefore, this study should be interpreted with caution about whether the results reflect the general Japanese public’s attitudes. Nevertheless, this study’s findings mirror existing perceptions among cancer patients towards the FCOI of physicians. Third, we did not collect data on the physicians who oversaw the participants’ treatments as cancer patients. Other unexamined confounding factors, such as physician specialty, details of their FCOI with Pharma, would influence the differences of participant awareness and perception of physician-Pharma interactions.[12, 24, 57] Despite the limitations, to the best of our knowledge, this is the first study to examine the awareness and perceptions of physician-Pharma interactions among non-healthcare professionals in Japan. The opinions of cancer patients and the supporting public could help explore appropriate FCOI management methods in Japan and other countries.

## Conclusion

While most participants were aware of some FCOI between healthcare sectors and Pharma in Japan, its extent differed much depending on the interactions. Participants also reported a significantly decreased trust in physicians who received personal gifts and payments than those accepting office-use related gifts. In addition, several participants expected that physicians should be highly ethical, minimize FCOI outside of research, and assume that FCOI disproportionately influences a physicians’ clinical practice, increases healthcare costs, and lowers patients’ trust in physicians. Further steps are required to improve patient awareness of FCOI, patient trust, and transparency in healthcare. These include improving public awareness with seminars focusing on direct physician-patient communication, more patient-oriented regulation of FCOI, and educating medical students and physicians about FCOI.

## Data Availability

The data that support the findings of this study are available on request from the corresponding author, AM. The data are not publicly available due to their containing information that could compromise the privacy of research participants.

## Acknowledgement

Authors appreciate Dr. Derek Hagman for professional English language editing and constructive opinions for this study.

## Author contributions

We describe contributions to the paper using the CRediT taxonomy provided above. Conceptualization: all authors;

Data curation: A.M.;

Formal analysis: A.M.;

Funding acquisition: A.O.;

Investigation: all authors; Methodology: all authors;

Project Administration: A.O., and T.T.;

Supervision: A.O., and T.T;

Visualization: A.M., A.O., and T.T.;

Writing – Original Draft: Y.S., A.M., K.H.;

Writing – review & editing: A.M., Y.K., T.T., and A.O.

All authors revised the manuscript critically for important intellectual content, gave final approval of the revision to be published, and agreed to be accountable for all the aspects of the study.

## Funding

This study was funded in part by the Medical Governance Research Institute. This non-profit enterprise receives donations from pharmaceutical companies, including Ain Pharmaciez, Inc., other organizations, and private individuals. This study also received support from the Tansa (formerly known as Waseda Chronicle), an independent non-profit news organization dedicated to investigative journalism. Ain Pharmacies had no role in the design and conduct of the study, the collection, management, analysis, and interpretation of the data, the preparation, review, and approval of the manuscript, or the decision to submit the manuscript for publication. Tansa was engaged in the collection and management of the payment data, but had no role in the design and conduct of the study, the analysis and interpretation of the data, the preparation, review and approval of the manuscript, or the decision to submit the manuscript for publication.

Supporting information:

## Supplementary Method

### Variables

#### Sociodemographic factors

Sociodemographic factors included age, sex, highest educational attainment (*less than high school graduate; high school graduate; associate degree or diploma; or bachelor’s degree or more*), employment status (*full time job; self-employed; part-time job; unemployed; or other*), and annual income (*<2 million JPY (<18,349 USD); 2-4 million JPY (18,349-36,697 USD); 4-6 million JPY (36,697-55,046 USD); 6-8 million JPY (55,046-73,394 USD); 8-10 million JPY (73,394-91,743 USD); or >10 million JPY (>91,743 USD*).

#### Primary diagnosis

We asked participant’s primary disease and categorized it into seven types: *non-cancer, prostate cancer, lung cancer, breast cancer, colorectal cancer, gastric cancer, and other kinds of cancer*.

*Clinical factors relating to cancer (only for cancer patients).* For cancer patients, we collected data on clinical characteristics including disease stage (*Stage I, Stage II, Stage III, Stage IV, or unclear*), year of diagnosis (*2018, 2017, 2016, 2015, or before 2015*), type of hospital in which participant received treatment (*Cancer hospital, national university hospital, private university hospital, national municipal hospital, private municipal hospital, or other hospital*), the treatment participants had been receiving (*no active treatment, anticancer drug, molecularly targeted drug, immune checkpoint inhibitor, hormonal therapy, radiation therapy, or other therapy*), treatments participants had ever received (*no active treatment, anticancer drug, molecularly targeted drug, immune checkpoint inhibitor, hormonal therapy, radiation therapy, surgery, or other therapy*), and disease recurrence (*yes, no, not clear, or other*).

#### Awareness of physician-Pharma interactions

To assess awareness of physician-Pharma interactions, we evaluated awareness of 11 types of interactions, including pamphlets and leaflets, stationeries, textbooks and articles, free drug samples, free meals travel and accommodations, lecture fees, disclosure program of the payment from Pharma to physicians, conduct of research for Pharma, outsourcing expenses, and possession of stocks. Consequently, we used the following question, *“Do you know that some physicians would receive pamphlet and leaflet concerning products manufactured by pharmaceutical companies from their sales representatives?”* on a three-point Likert scale (*Yes, No, or Not sure*). Other items are listed in Supplementary Material 2.

#### Influence on trust in physicians

We assessed the impact on trust in physicians over 11 types of interactions including, pamphlets and leaflets, stationeries, textbooks and articles, free drug samples, free meals, travel and accommodations, lecture fees, conduct of research for Pharma, outsourcing expenses, possession of stocks, and insider trading. Consequently, we used the following question, *“How would your trust in your physician be influenced if your physician received pamphlet or leaflet with information about the products from a pharmaceutical sales representative?”* on a five-point Likert scale (*Increase of trust in physicians, Slight increase of trust in physicians, Neither increase nor decrease of trust in physicians, Slight decrease of trust in physicians, or Decrease of trust in physicians*). Other items are listed in Supplementary Material 3.

#### Perception on physician-Pharma interaction

We evaluated participants’ perception of physician-Pharma interactions by assessing whether to agree with the following statements. 1) *“Payments from pharmaceutical companies would influence physician prescription pattern.”* 2) *“Payments from pharmaceutical companies would increase unnecessary prescriptions and healthcare costs.”* 3) *“A receipt of payments from pharmaceutical companies would be acceptable as long as its amount is small.”* 4) *“Physician’s receipts of gifts, meals and honoraria from pharmaceutical companies are unethical.”* 5) *“Physician’s receipts of gifts, meals, and honoraria from pharmaceutical companies would undermine the trust in physicians.”* 6) *“More rigorous internal regulation of gifts, meals, and honoraria from pharmaceutical companies to physicians is needed among pharmaceutical companies.”* 7) *“More rigorous internal regulation of gifts, meals, and honoraria from pharmaceutical companies to physicians is needed among physicians.”* 8) *“More rigorous legal regulation of gifts, meals, and honoraria from pharmaceutical companies to physicians is needed.”* On a five-point Likert scale (*Agree, Slightly agree, Neither agree nor disagree, Slightly disagree, or Disagree*). Further, for participants who agree or slightly agree with the third statement, we additionally asked the closet amount of money which they think is acceptable on a four-point scale (*1,000 JPY (9.2 USD) or below, 1,000-3,000 JPY (9.2-27.5 USD), 3,000-10,000 JPY (27.5-91.7 USD), or other*). For participants who agreed or slightly agreed with either of sixth, seventh or eighth statement, we additionally asked the monetary value (*100 thousand JPY (917 USD) or below, 100 thousand-1 million JPY (917-9174 USD), 1-5 million JPY (9,174-45,872 USD), 5-10 million JPY (45,872-91,743 USD, or 10-30 million JPY (91,743-275,229 USD)*) and frequency (*twice a week, once a week, once every two weeks, once a month, or once every few months*) of the financial interaction with the Pharma. (Supplementary Material 4)

#### Attitude towards various professional FCOI

Attitude towards various professional FCOI was assessed by using the following five statements. 1) *“It is problematic for judges to receive gifts, meals, and other entertainment from the trial attorney in charge.”* 2) *“It is problematic for sports referees to receive gifts, meals, and other entertainment from the players of the games they are assigned to.”* 3) *“It is problematic for politicians to receive gifts, meals, and other entertainment from lobbyists.”* 4) *“It is problematic for physicians to receive gifts, meals, and other entertainment from pharmaceutical sales representatives.”* 5) *“It is problematic for business people to receive gifts, meals, and other entertainment from their customers.”* On a five-point Likert scale (*Agree, Slightly agree, Neither agree nor disagree, Slightly disagree, or Disagree*).

*An open-ended question about participants’ perception of FCOI between Pharma and physicians.* To dig deeper into participants’ perceptions on FCOI between Pharma and physicians, we investigated responses to an open-ended question: “*Please freely describe your thoughts about non-research-related offerings from Pharma to a physician (e.g., gifts, free meals, monetary incentives for a lecture).*” The responses to this question were analyzed separately through qualitative analysis.

**Supplementary Material 2. List of questions regarding awareness of physician-Pharma interactions.**

**Supplementary Material 3. List of questions regarding influence on trust in physicians.**

**Supplementary Material 4. List of questions regarding perception on physician-Pharma interactions.**

**Supplementary Material 5. Demographic breakdown on the number and percent of participants who were aware of at least one physician-Pharma interaction.**

^1^The analysis included only participants with cancer. Abbreviations: JPY; Japanese yen, USD; US dollar, Pharma; Pharmaceutical Company.

**Supplementary Material 6. Logistic regression analysis of participants’ awareness of physician-Pharma interactions.**

^1^The analysis included only participants with cancer. Abbreviations: JPY; Japanese yen, Pharma; Pharmaceutical Company, Ref; reference value.

**Supplementary Material 7. Number and percent of participants who reported decreased trust in at least one physician-Pharma interaction by participant demographics.**

^1^The analysis included only participants with cancer. Abbreviations: JPY; Japanese yen, Pharma; Pharmaceutical Company.

**Supplementary Material 8. Logistic regression analysis of the influence of physician-Pharma relationships on participants’ trust.**

^1^ The analysis included only participants with cancer. There were no statistically significant differences between the influence of physician-pharmaceutical company relationships on participants’ trust and each variable. Abbreviations: JPY; Japanese yen, Pharma; Pharmaceutical Company.

