## Supplementary Material 1-8 for "Awareness and perceptions among members of a Japanese cancer patient advocacy group concerning the financial relationships between the pharmaceutical industry and physicians: a mixed-methods analysis of survey data"

**Supplementary Material 2. List of questions regarding awareness of physician-Pharma interactions.**

|  | Question | Response options |
| --- | --- | --- |
| 1 | Do you know that some physicians would receive pamphlet and leaflet concerning products manufactured by pharmaceutical companies from their sales representatives? | Yes  No  Not sure |
| 2 | Do you know that some physicians would receive stationeries embedded with pharmaceutical company names from pharmaceutical companies? | Yes  No  Not sure |
| 3 | Do you know that some physicians would receive textbooks and academic articles for daily practice from pharmaceutical companies? | Yes  No  Not sure |
| 4 | Do you know that some physicians would receive free drug samples from pharmaceutical companies? | Yes  No  Not sure |
| 5 | Do you know that some physicians would receive free meals from pharmaceutical companies at study meetings and conferences? | Yes  No  Not sure |
| 6 | Do you know that some physicians would receive travel and accommodation reimbursements from pharmaceutical companies to attend conferences? | Yes  No  Not sure |
| 7 | Do you know that some physicians would receive 10-100 thousand JPY (about 100-1000 USD) as honoraria from pharmaceutical companies when they serve as lecturers at conferences held by pharmaceutical companies? | Yes  No  Not sure |
| 8 | Do you know that pharmaceutical companies disclose payments made to healthcare professionals and healthcare organizations for promotional and other purposes? | Yes  No  Not sure |
| 9 | Do you know that some physicians would participate in researches sponsored by pharmaceutical companies? | Yes  No  Not sure |
| 10 | Do you know that some healthcare organizations receive several hundred thousand JPY of honoraria from pharmaceutical companies in exchange for enrolment of their patients for treatment in clinical trials sponsored by pharmaceutical companies? | Yes  No  Not sure |
| 11 | Do you know some physicians would have stock in pharmaceutical companies? | Yes  No  Not sure |

**Supplementary Material 3. List of questions regarding influence on trust in physicians.**

|  | Question | Response options |
| --- | --- | --- |
| 1 | How would your trust in your physician be influenced if your physician received pamphlets or leaflets with information about the products from a pharmaceutical sales representative? | Increase in trust of physicians  Slight increase in trust of physicians  Neither increase nor decrease in trust of physicians  Slight decrease in trust of physicians  Decrease in trust of physicians |
| 2 | How would your trust in your physician be influenced if your physician received stationeries with a pharmaceutical company name from pharmaceutical companies? | Increase in trust of physicians  Slight increase in trust of physicians  Neither increase nor decrease in trust of physicians  Slight decrease in trust of physicians  Decrease in trust of physicians |
| 3 | How would your trust in your physician be influenced if your physician received textbooks or articles concerning daily clinical practice from pharmaceutical companies? | Increase in trust of physicians  Slight increase in trust of physicians  Neither increase nor decrease in trust of physicians  Slight decrease in trust of physicians  Decrease in trust of physicians |
| 4 | How would your trust in your physician be influenced if your physician received free drug samples from pharmaceutical companies? | Increase in trust of physicians  Slight increase in trust of physicians  Neither increase nor decrease in trust of physicians  Slight decrease in trust of physicians  Decrease in trust of physicians |
| 5 | How would your trust in your physician be influenced if your physician received free meals from pharmaceutical companies at training sessions and lecture meetings? | Increase in trust of physicians  Slight increase in trust of physicians  Neither increase nor decrease in trust of physicians  Slight decrease in trust of physicians  Decrease in trust of physicians |
| 6 | How would your trust in your physician be influenced if your physician received travel and accommodation reimbursement from pharmaceutical companies to attend lecture meetings? | Increase in trust of physicians  Slight increase in trust of physicians  Neither increase nor decrease in trust of physicians  Slight decrease in trust of physicians  Decrease in trust of physicians |
| 7 | How would your trust in your physician be influenced if your physician received 10-100 thousand JPY (about 100-1000 USD) as lecture fees from pharmaceutical companies for lecturing about its drugs on behalf of pharmaceutical companies at meetings? | Increase in trust of physicians  Slight increase in trust of physicians  Neither increase nor decrease in trust of physicians  Slight decrease in trust of physicians  Decrease in trust of physicians |
| 8 | How would your trust in your physician be influenced if your physician conducted research sponsored by pharmaceutical companies? | Increase in trust of physicians  Slight increase in trust of physicians  Neither increase nor decrease in trust of physicians  Slight decrease in trust of physicians  Decrease in trust of physicians |
| 9 | How would your trust in your physician be influenced if your healthcare organization received several hundred thousand JPY honoraria from pharmaceutical companies in exchange for enrolment of their patients with treatment in a clinical trial sponsored by a pharmaceutical company? | Increase in trust of physicians  Slight increase in trust of physicians  Neither increase nor decrease in trust of physicians  Slight decrease in trust of physicians  Decrease in trust of physicians |
| 10 | How would your trust in your physician be influenced if your physician had stock in pharmaceutical companies? | Increase in trust of physicians  Slight increase in trust of physicians  Neither increase nor decrease in trust of physicians  Slight decrease in trust of physicians  Decrease in trust of physicians |
| 11 | How would your trust in your physician be influenced if your physician knew the promising new drug before the information became public and purchased the stock in the pharmaceutical company? | Increase in trust of physicians  Slight increase in trust of physicians  Neither increase nor decrease in trust of physicians  Slight decrease in trust of physicians  Decrease in trust of physicians |

**Supplementary Material 4. List of questions regarding perception on physician-Pharma interactions.**

|  | Question | Response options |
| --- | --- | --- |
| 1 | Payments from pharmaceutical companies would influence physician prescription patterns. | Agree  Slightly agree  Neither agree nor disagree  Slightly disagree  Disagree |
| 2 | Payments from pharmaceutical companies would increase unnecessary prescriptions and healthcare costs. | Agree  Slightly agree  Neither agree nor disagree  Slightly disagree  Disagree |
| 3 | A receipt of payments from pharmaceutical companies would be acceptable as long as its amount is small. | Agree  Slightly agree  Neither agree nor disagree  Slightly disagree  Disagree |
| 4 | Physician’s receipts of gifts, meals, and honoraria from pharmaceutical companies are unethical. | Agree  Slightly agree  Neither agree nor disagree  Slightly disagree  Disagree |
| 5 | Physician’s receipts of gifts, meals, and honoraria from pharmaceutical companies would undermine the trust in physicians. | Agree  Slightly agree  Neither agree nor disagree  Slightly disagree  Disagree |
| 6 | More rigorous internal regulation of gifts, meals, and honoraria from pharmaceutical companies to physicians is needed among pharmaceutical companies. | Agree  Slightly agree  Neither agree nor disagree  Slightly disagree  Disagree |
| 7 | More rigorous internal regulation of gifts, meals, and honoraria from pharmaceutical companies to physicians is needed among physicians. | Agree  Slightly agree  Neither agree nor disagree  Slightly disagree  Disagree |
| 8 | More rigorous legal regulation of gifts, meals, and honoraria from pharmaceutical companies to physicians is needed. | Agree  Slightly agree  Neither agree nor disagree  Slightly disagree  Disagree |
| 9 | For participants who agreed or slightly agreed with the third statement, what is the closest amount of money that you consider as a small amount? | 1,000 JPY or below  1,000-3,000 JPY  3,000-10,000 JPY  Other |
| 10 | For participants who agreed or slightly agreed with either the sixth, seventh or eighth statements, what is the closest thing to the frequency of non-research benefits from pharmaceutical companies to physicians that you consider acceptable? | 100 thousand JPY or below  100 thousand-1 million JPY  1-5 million JPY  5-10 million JPY  10-30 million JPY |
| 11 | For participants who agreed or slightly agreed with either the sixth, seventh or eighth statements, what is the closest approximation to the annual total amount of non-research benefits from pharmaceutical companies to physicians that you consider to be acceptable? | Twice a week  Once a week  Once every two weeks  Once a month  Once every few months |

**Supplementary Material 5. Demographic breakdown on the number and percent of participants who were aware of at least one physician-Pharma interaction.**

| Variables | Number (%) | | *P* value |
| --- | --- | --- | --- |
|  | **Aware** | **Unaware or not sure** |  |
| Gender |  |  |  |
| Male | 49 (77.8) | 14 (22.2) | 0.053 |
| Female | 28 (93.3) | 2 (6.7) |  |
| Age category |  |  |  |
| ≦60 | 13 (86.7) | 2 (13.3) | 0.879 |
| 61-70 | 24 (80.0) | 6 (20.0) |  |
| ≥71 | 39 (79.6) | 10 (20.4) |  |
| Income |  |  |  |
| Lower income (<4 million JPY (<36,697 USD)) | 39 (84.8) | 7 (15.2) | 0.129 |
| Higher income (≥4 million JPY (≥36,697 USD)) | 29 (72.5) | 11 (27.5) |  |
| Job |  |  |  |
| Employed | 30 (81.1) | 7 (18.9) | 0.576 |
| Unemployed | 45 (80.4) | 11 (19.6) |  |
| Education |  |  |  |
| High school graduate or less | 21 (75.0) | 7 (25.0) | 0.264 |
| Associate degree or more | 54 (83.1) | 11 (16.9) |  |
| Cancer |  |  |  |
| Non-cancer participants | 17 (85.0) | 3 (15.0) | 0.433 |
| Cancer patients | 55 (79.7) | 14 (20.3) |  |
| Cancer stage^1^ |  |  |  |
| 1 | 17 (73.9) | 6 (26.1) | 0.287 |
| 2-4 | 26 (83.9) | 5 (16.1) |  |
| Year^1^ |  |  |  |
| 2015-2018 | 15 (88.2) | 2 (11.8) | 0.320 |
| Before 2015 | 41 (78.9) | 11 (21.1) |  |
| Hospital^1^ |  |  |  |
| Other hospitals | 24 (80.0) | 6 (20.0) | 0.555 |
| Cancer special hospitals | 31 (81.6) | 7 (18.4) |  |
| Previous cancer recurrence^1^ |  |  |  |
| No or other | 41 (78.9) | 11 (21.1) | 0.246 |
| Yes | 12 (92.3) | 1 (7.7) |  |
| Experience with pharmacotherapy^1^ |  |  |  |
| No | 19 (73.1) | 7 (26.9) | 0.145 |
| Yes | 33 (86.8) | 5 (13.2) |  |
| Experience with radiotherapy^1^ |  |  |  |
| No | 23 (85.2) | 4 (14.8) | 0.447 |
| Yes | 29 (80.6) | 7 (19.4) |  |
| Previous surgical treatment^1^ |  |  |  |
| No | 22 (75.9) | 7 (25.1) | 0.292 |
| Yes | 32 (84.2) | 6 (15.8) |  |

^1^The analysis included only participants with cancer. Abbreviations: JPY; Japanese yen, USD; US dollar, Pharma; Pharmaceutical Company.

**Supplementary Material 6. Logistic regression analysis of participants' awareness of physician-Pharma interactions.**

| Variables | Odds ratio (95% confidence interval) | *P* value |
| --- | --- | --- |
| Gender |  |  |
| Male | Ref. |  |
| Female | 4.00 (0.85–18.90) | 0.080 |
| Age category |  |  |
| ≦60 | Ref. |  |
| 61-70 | 0.62 (0.11–3.49) | 0.584 |
| ≥71 | 0.60 (0.12–3.10) | 0.542 |
| Income |  |  |
| Lower income  (<4 million JPY) | Ref. |  |
| Higher income  (≥4 million JPY) | 0.47 (0.16–1.37) | 0.168 |
| Job |  |  |
| Employed | Ref. |  |
| Unemployed | 0.95 (0.33–2.74) | 0.931 |
| Education |  |  |
| High school graduate or less | Ref. |  |
| Associate degree or more | 1.64 (0.56–4.79) | 0.369 |
| Cancer |  |  |
| Non-cancer participants | Ref. |  |
| Cancer patients | 0.69 (0.18–2.70) | 0.598 |
| Cancer stage^1^ |  |  |
| 1 | Ref. |  |
| 2-4 | 1.84 (0.48–6.97) | 0.373 |
| Year^1^ |  |  |
| 2015-2018 | Ref. |  |
| Before 2015 | 0.50 (0.10–2.51) | 0.397 |
| Hospital^1^ |  |  |
| Other hospitals | Ref. |  |
| Cancer special hospitals | 1.11 (0.33–3.73) | 0.869 |
| Previous cancer recurrence^1^ |  |  |
| No or other | Ref. |  |
| Yes | 3.22 (0.38–27.52) | 0.286 |
| Experience with pharmacotherapy^1^ |  |  |
| No | Ref. |  |
| Yes | 2.43 (0.68–8.74) | 0.173 |
| Experience with radiotherapy^1^ |  |  |
| No | Ref. |  |
| Yes | 0.72 (0.19–2.76) | 0.633 |
| Previous surgical treatment^1^ |  |  |
| No | Ref. |  |
| Yes | 1.70 (0.50–5.74) | 0.395 |

^1^The analysis included only participants with cancer. Abbreviations: JPY; Japanese yen, Pharma; Pharmaceutical Company, Ref; reference value.

**Supplementary Material 7. Number and percent of participants who reported decreased trust in at least one physician-Pharma interaction by participant demographics.**

| Variables | Number (%) | | *P* value |
| --- | --- | --- | --- |
|  | Decrease trust | Other |  |
| Gender |  |  |  |
| Male | 55 (87.3) | 8 (12.7) | 0.157 |
| Female | 28 (96.6) | 1 (3.4) |  |
| Age category |  |  |  |
| ≦60 | 14 (93.3) | 1 (6.7) | 0.550 |
| 61-70 | 26 (89.7) | 3 (10.3) |  |
| ≥71 | 44 (89.8) | 5 (10.2) |  |
| Income |  |  |  |
| Lower income (<4 million JPY) | 41 (89.1) | 5 (10.9) | 0.590 |
| Higher income (≥4 million JPY) | 36 (90.0) | 4 (10.0) |  |
| Job |  |  |  |
| Employed | 33 (98.2) | 4 (10.8) | 0.515 |
| Unemployed | 51 (91.1) | 5 (8.9) |  |
| Education |  |  |  |
| High school graduate or less | 23 (82.1) | 5 (17.7) | 0.089 |
| Associate degree or more | 61 (93.9) | 4 (6.1) |  |
| Cancer |  |  |  |
| Non-cancer participants | 19 (95.0) | 1 (5.0) | 0.424 |
| Cancer patients | 62 (89.9) | 7 (10.1) |  |
| Cancer stage^1^ |  |  |  |
| 1 | 22 (95.7) | 1 (4.3) | 0.283 |
| 2-4 | 27 (87.1) | 4 (12.9) |  |
| Year^1^ |  |  |  |
| 2015-2018 | 14 (82.4) | 3 (17.6) | 0.316 |
| Before 2015 | 46 (90.2) | 5 (9.8) |  |
| Hospital^1^ |  |  |  |
| Other hospitals | 26 (89.7) | 3 (10.3) | 0.517 |
| Cancer special hospitals | 33 (86.8) | 5 (13.2) |  |
| Previous cancer recurrence^1^ |  |  |  |
| No or other | 47 (90.4) | 5 (9.6) | 0.428 |
| Yes | 11 (84.6) | 2 (15.4) |  |
| Experience with pharmacotherapy^1^ |  |  |  |
| No | 24 (92.3) | 2 (7.7) | 0.398 |
| Yes | 33 (86.8) | 5 (13.2) |  |
| Experience with radiotherapy^1^ |  |  |  |
| No | 25 (92.6) | 2 (7.4) | 0.349 |
| Yes | 31 (86.1) | 5 (13.9) |  |
| Previous surgical treatment^1^ |  |  |  |
| No | 24 (82.8) | 5 (17.2) | 0.119 |
| Yes | 36 (94.7) | 2 (5.3) |  |

^1^The analysis included only participants with cancer. Abbreviations: JPY; Japanese yen, Pharma; Pharmaceutical Company.

**Supplementary Material 8. Logistic regression analysis of the influence of physician-Pharma relationships on participants' trust.**

| Variables | Odds ratio (95% confidence interval) | *P* value |
| --- | --- | --- |
| Gender |  |  |
| Male | Ref. |  |
| Female | 4.07 (0.48–34.21) | 0.185 |
| Age category |  |  |
| ≦60 | Ref. |  |
| 61-70 | 0.62 (0.059–6.52) | 0.713 |
| ≥71 | 0.63 (0.068–5.84) | 0.683 |
| Income |  |  |
| Lower income  (<4 million JPY) | Ref. |  |
| Higher income  (≥4 million JPY) | 1.10 (0.27–4.40) | 0.895 |
| Job |  |  |
| Employed | Ref. |  |
| Unemployed | 1.24 (0.31–4.94) | 0.764 |
| Education |  |  |
| High school graduate or less | Ref. |  |
| Associate degree or more | 3.32 (0.82–13.44) | 0.093 |
| Cancer |  |  |
| Non-cancer participants | Ref. |  |
| Cancer patients | 0.47 (0.054–4.03) | 0.488 |
| Cancer stage^1^ |  |  |
| 1 | Ref. |  |
| 2-4 | 0.31 (0.032–2.95) | 0.306 |
| Year^1^ |  |  |
| 2015-2018 | Ref. |  |
| Before 2015 | 1.97 (0.42–9.30) | 0.376 |
| Hospital^1^ |  |  |
| Other hospitals | Ref. |  |
| Cancer special hospitals | 0.76 (0.17–3.48) | 0.689 |
| Previous cancer recurrence^1^ |  |  |
| No or other | Ref. |  |
| Yes | 0.59 (0.10-3.42) | 0.552 |
| Experience with pharmacotherapy^1^ |  |  |
| No | Ref. |  |
| Yes | 0.55 (0.098–3.08) | 0.496 |
| Experience with radiotherapy^1^ |  |  |
| No | Ref. |  |
| Yes | 0.50 (0.089–2.78) | 0.425 |
| Previous surgical treatment^1^ |  |  |
| No | Ref. |  |
| Yes | 3.75 (0.67–20.93) | 0.132 |

^1^ The analysis included only participants with cancer. There were no statistically significant differences between the influence of physician-pharmaceutical company relationships on participants' trust and each variable. Abbreviations: JPY; Japanese yen, Pharma; Pharmaceutical Company.
